# International Journal of Epidemiology peer reviewers: global and gender distribution

**DOI:** 10.1101/2020.05.14.20101964

**Authors:** Gwinyai Masukume, Julio González-Álvarez, Victor Grech

## Abstract

The science of epidemiology studies and analyses the patterns, distributions and determinants of health-related entities. Peer review is crucial to the biomedical journal ecosystem. Furthermore, gender inequality persists in science and improving gender representation is a key international goal. For these reasons, we evaluated the geographical location and gender distribution of the *International Journal of Epidemiology* peer reviewers for 2018.

There were 695 peer reviewers hailing from 41 countries. 77% (533) of the reviewers were from 20% (8) of these countries, a classical 80:20 Pareto distrubution.

41% (282) of peer reviewers were female. Although in line with the general underrepresentation of females reported in the literature, this proportion was much closer to parity than in other fields.

Our investigation of peer reviewer ‘epidemiology’ demonstrates geographical and gender inequalities that can be further improved since diversity promotes innovation and a greater possibility for solving complex problems.

---

The science of epidemiology studies and analyses the patterns, distributions and determinants of health-related entities.^[1]^ The *International Journal of Epidemiology* (*IJE*) content is a valuable resource that keeps readers abreast of the latest advances in this field.^[2]^ Peer reviewers are vital to the biomedical journal ecosystem.^[3]^ We studied and analysed the ‘epidemiology’ of *IJE* peer reviewers using 2018 data (Figure 1).^[4]^

**Figure 1.**
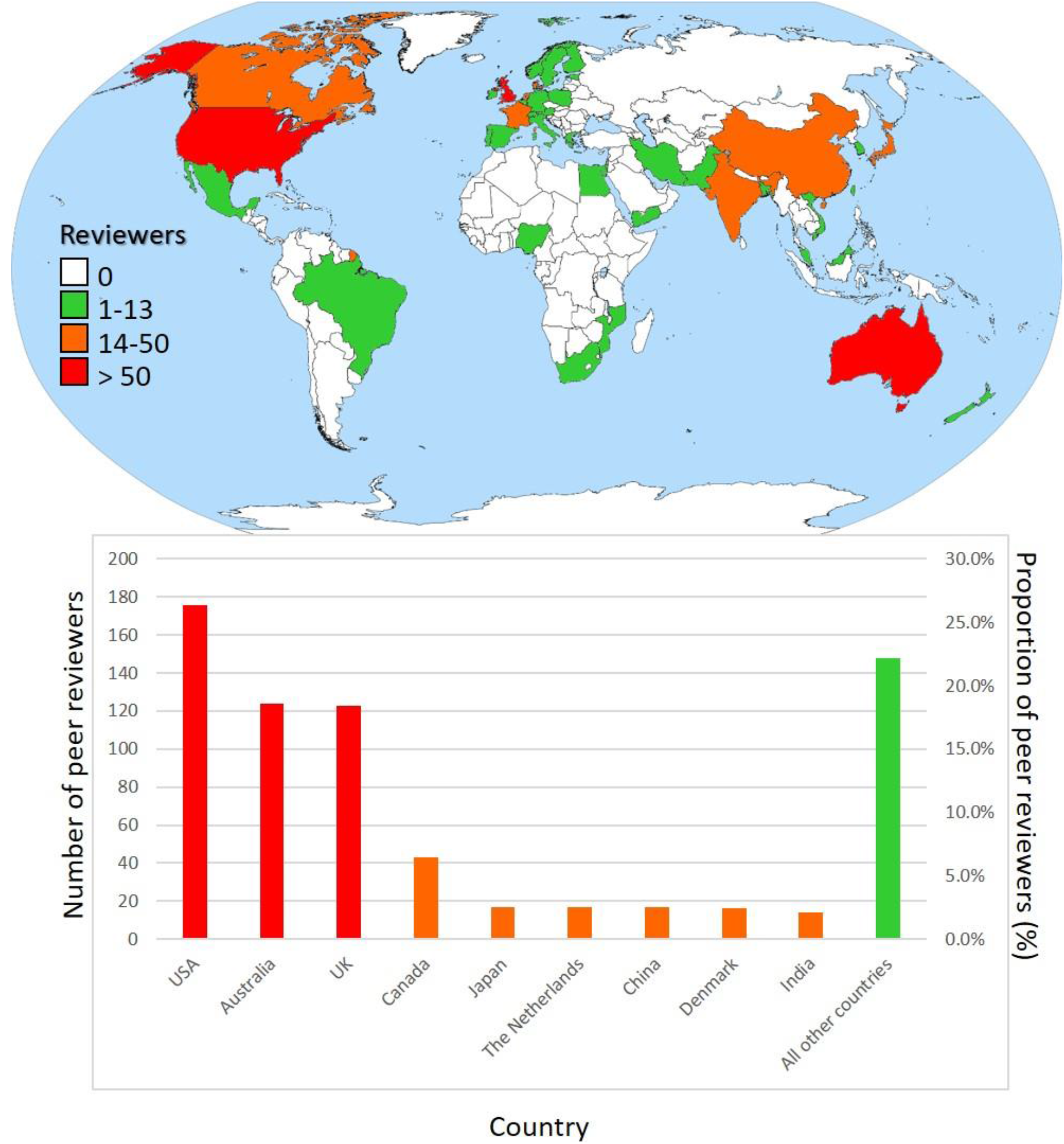
Number and location of *The International Journal of Epidemiology* peer reviewers, 2018.

There were 695 peer reviewers from all six permanentely inhabitated continents, hailing from 41 countries. 77% (533) of the reviewers were from 20% (8) of these countries. This pattern adheres to the 80/20 Pareto principle (80% of events emanate from 20% of attributary causes). Ranking was: South America 1% (8), Africa 2% (13), Asia 10% (69), Oceania 19% (130), North America 32% (222) and Europe 36% (253).

A previous analysis of *IJE* authors revealed that among first authors, 44% of them were from the United States of America (USA) and United Kingdom (UK). Our findings mirror these with 43% (299) of the reviewers from the USA and UK. Rounding out the top three referee countries was Australia with 18% (124). This top three country distribution was similar to that of *The Lancet’s*, another leading medical journal.^[5]^

The greater proportion of referees from high-income countries is determined by availability of more research funding, better access to reliable census and mortality data, more possibilty of protected time to do research and greater facility with the English language.^[4]^

Because gender inequality exists in science, we also considered peer reviewer gender.^[6]^ We used https://gender-api.com/, the largest online resource that determines gender based on first name. This was supplemented by ascertaining gender from internet searches for reviewer biographical and photographic information. We found that 41% (282) of peer reviewers were female. Although in line with the general underrepresentation of females reported in the literature, this proportion was much closer to parity than in other fields.^[6]^ Promoting gender equity is a key aspect of the international Sustainable Development Goals.^[7]^

It is worth noting that one reviewer could have performed multiple reviews and our analysis did not take this into account. In addition, reviewer numbers were absolute counts and not relative to a country’s population.

Our concise investigation of *IJE* peer reviewer ‘epidemiology’ allows a baseline from which geographical and gender representation can be further improved. Greater diversity promotes innovation and a higher possibility for solving complex problems.^[8]^

## Data Availability

The dataset is publicly available.

## Ethics

Research ethics committee approval was not required because of the public nature of the dataset.

## References

1. Anderson W. The history in epidemiology. International journal of epidemiology. 2018;48(3):672–4.

2. Grant C, Williams B, Driscoll T. Historical trends in publications in the International Journal of Epidemiology. International journal of epidemiology. 2018;47(3):938–41.

3. Glonti K, Boutron I, Moher D, Hren D. Journal editors’ perspectives on the roles and tasks of peer reviewers in biomedical journals: a qualitative study. BMJ Open. 2019;9(11):e033421.

4. Leeder S. The IJE and the volatile world of academic publication. International journal of epidemiology. 2019;48(2):323–31.

5. Masukume G, Grech V. The Lancet peer reviewers: global pattern and distribution. Lancet (London, England). 2018;391(10140):2603–4.

6. González-Alvarez J, Cervera-Crespo T. Psychiatry research and gender diversity: authors, editors, and peer reviewers. Lancet Psychiatry. 2019;6(3):200–1.

7. Gupta GR, Oomman N, Grown C, Conn K, Hawkes S, Shawar YR, et al. Gender equality and gender norms: framing the opportunities for health. Lancet (London, England). 2019;393(10190):2550–62.

8. Phillips KW. How Diversity Works. Scientific American. 2014;311(4):42–7.

